# The level of liver and renal function biomarker abnormalities among hospitalized COVID-19 patients in Ethiopia

**DOI:** 10.1101/2022.02.15.22271010

**Authors:** Yakob Gebregziabher Tsegay, Molalegne Bitew, Tigist Workneh, Assegdew Atlaw, Mintsnot Aragaw, Mesay Gemechu, Nega Brhane

## Abstract

**Background:** COVID-19 pandemic is unprecedented public health emergency and added burden to developing countries. The pandemic cause multi organ failures (MOF) predominantly affects lung, cardiac, renal and liver organs as severity of the disease exacerbates. That is the rationale to execute this study with the aim to determine the magnitude of abnormal organ function test parameters and its association between markers of organ failure and disease severity in patients infected with COVID-19 admitted at Millennium COVID-19 Care Center (CCC).

**Methods:** A cross-sectional study was conducted among COVID-19 patients admitted at Millennium COVID-19 Care and Treatment Center (MCCTC) from May 2021 up to Oct 2021. In this study 500 participant’s information were collected from the laboratory database of Millennium COVID-19 care center. Data were analyzed using SPSS version 25. P-value <0.05 was considered significantly associated.

**Result:** The median age of the 500 study participants was 55.6±7.7 years, and from these 67.6% of patients were males. Liver function parameters Aspartae transferase (AST),) alanine aminotransferase (ALT) and Alakaline phosphatase (ALP) the mean value of overall patients were elevated and three of these parameters were highly elevated among critical patients (56.9±57.7, 58.5±6, and 114.6±6) respectively. All study participants had an elevated Creatinine. 66.8% males, 65% Intensive care unit (ICU), had an elevated serum value of ALT and AST respectively. Troponin was found elevated among males (54%) and 59% among ICU (critical) patients.

**Conclusion:** Liver and renal function test biomarkers such as creatine kinase muscle-brain isoenzymes (CK-MB), troponin, AST, ALT and Creatinine serum value was found elevated among ICU than non ICU patients. Organ function biomarkers are a candidate for predicting COVID-19 disease severity in order to guide clinical care.

## Introduction

SARS COV-2 primarily causes pneumonia and the disease was named as novel coronavirus acute respiratory distress (ARS) (COVID-19)[1, 2]. COVID-19 symptoms are highly variable, ranging from severe illness to none. It can spread mainly through the air when people are in close contact or near to each other. Human-to-human transmission is the main way of cause infection and it could also spread via contaminated surfaces [3, 4]. People remain infectious for up to two weeks, and can spread the virus even if they do not show symptoms [5].

The corona virus pandemic is a global health crisis of our time and the greatest challenges ever have faced since its emergence in Wuhan, China. It has become a pandemic that has heavily affected the global population [6-8]. As of November 18, 2021, there have been 256,402,576 confirmed cases of COVID-19 and 5,148,538 deaths, reported to World Health organization (WHO). Similarly, there have been 369,667 confirmed cases of COVID-19 with 6,655 deaths in Ethiopia[9]. Severe Acute Respiratory Distress corona Virus (SARS COV-2) infection is characterized by mild symptoms for most patients but for some individuals requires admission at intensive care [10, 11].

COVID-19 is characterized by acute respiratory failure and diffuse alveolar damage at lung [12], the involvement of other organs was also mentioned by different authors [13-16]. After SARS COV-2 infects the lung, the virus may migrate to the blood, accumulate in kidney, and cause damage to resident renal cells [17, 18]. Reports showed that 6.7 to 11.4% of patients with SARS COV-2 results acute kidney injury and the mortality of those with acute kidney injury was 80% to 91.7% [19-21]. During COVID-19 infection, multiple organs can be affected depending on the state and severity of the infection. An increased level of total bilirubin, Aspartae transferase (AST),) alanine aminotransferase (ALT) and Alkaline phosphatase (ALP) in blood indicates hepatocyte damage causes liver failure [22, 23].

COVID-19 could also cause heart muscle injury as determined by release of high-sensitivity troponin [3, 24]. The occurrences of high cardiac-specific structural protein in the serum are associated with severe disease progression. In addition to this blood clotting can also be impaired as result of systemic inflammation from the infection [24-26].

Vast information was obtained on COVID-19 clinical features, but limited information has been provided on organ function laboratory biomarker abnormalities and, especially, on the association of disease severity and organ function tests parameters in Ethiopia and in African context. Therefore, the aim of this study was to determine the magnitude of abnormal organ function test parameters in hospitalized patients with confirmed SARS COV-2 infection and to define the association between markers of organ failure and disease severity in patients infected with SARS COV-2 in Ethiopia.

## Materials and Methods

### Study design

It is Millennium COVID-19 Care and Treatment Center based observational and cross sectional study on patients who are tested positive for SARS-CoV-2.

### Sample size

The study collected 500 participant’s information on convenient basis from laboratory database of Millennium COVID-19 care center from May to Oct 2021. Requests to the laboratory are generated online, and the laboratory results are sent electronically from the laboratory information system (LIS) to the patient’s electronic medical record.

### Sample collection for clinical chemistry tests analysis

Patients who had been tested for liver function test, renal function test and cardiac marker tests were included in this laboratory based clinical data report from the patient request paper. For this purpose, 8ml of venous blood samples were collected by serum separating tubes (SST) for clinical chemistry test parameters. The liver function tests, renal function and cardiac markers were analyzed using COBAS 6000 automated clinical chemistry analyzer. In most COVID-19 infection, real-time reverse transcription polymerase chain reaction (qRT-PCR) test confirmed cases; routine clinical chemistry tests were performed to assess renal, cardiac and liver functions. Among these organ function tests, Alanine transaminase (ALT), Alkaline phosphatase (ALP) and Aspartate transaminase (AST) were included under liver function tests; whereas creatine kinase muscle-brain isoenzymes (CK-MB), troponin, and AST were included under cardiac function tests. In addition, creatinine and urea were mentioned as markers for renal function tests. All clinical laboratory tests and interpretation were done following the manufacturers’ recommendation and standard operating procedure. Liver test abnormalities were defined as the elevation of the following liver enzymes in serum: ALT >33 U/L, AST >35 U/L and alkaline phosphatase > 87 U/L. Renal test abnormalities were defined as the elevation of Creatinine (0.5-0.9mg/dl) and Urea (10-45mg/dl). Cardiac biomarker abnormalities were defined as the elevation of troponin (0-14pg/ml) and CKMB (0-25 U/L).

### Principle and procedure of COBAS 6000 clinical chemistry Analyzer

The COBAS 6000 analyzer is an automated, software-controlled system for clinical chemistry analysis with three main principles of photometric assay, Ion Selective electrode measurements and chemiloillunision. A photometric principle system measures the absorbance of the reaction mixture in reaction cells on the reaction disk is directly proportional to the concentration of the analyte. This system uses serum sample and performs the following analytes like AST, ALT, DBIL, BILT, CREJ, UREA, GLUC, TP, CHOL, TRIG, AMLYL, CK-MB, LDH-c, HDL-c, LIP, ALB, AMY-P, UA, PHOS, Mg++, and Ca++ etc. An ion-selective electrode measuring system measures the electromotive force (EMF) in millivolts between the electrode in the diluted sample solution and the electrode in the reference solution. The electrodes are selective for CL-, K+, and Na+ only. It uses serum/plasma, urine, CSF and supernatant sample types. It is designed for both quantitative and qualitative in vitro determinations using a large variety of tests for analysis.

### SARS-CoV-2 RNA extraction

The swab samples were collected according to the standard safety rules and protocols for SARS-CoV-2. The RNA was extracted with the QIAamp viral RNA mini kit (Qiagen #52,906) according to the standard kit protocol.

### Primers and Probes for laboratory COVID-19 detection

SARS-CoV-2 was confirmed using qRT-PCR-. Two pairs of primers targeting nucleo-capsid protein (N) and open reading frame 1ab (ORF1ab) were used to amplify and examine. The corresponding primer sequences for the amplification of N gene were Forward primer 5′-GGGGAACTTCTCCTGCTAGAAT-3’, reverse primer 5′-CAGACATTTTGCTCTCAAGCTG-3′, and the probe of the following sequence 5′-FAM-TTGCTGCTGCTTGACAGATT-TAMRA-3′. For ORF1ab gene forward primer with the sequence of 5′ CCCTGTGGGTTTTACACTTAA-3′ (F), reverse primer 5′ ACGATTGTGCATCAGCTGA-3′, and the probe with the following sequence 5′-CY3 CCGTCTGCGGTATGTGGAAAGGTTATGG-BHQ1-3′. These diagnostic criteria were based on the recommendations by the WHO. The SARS-CoV-2 RNA detection was done according to the standard kit protocol used by the MCCC.

### Operational case definitions

#### Moderate cases

According to WHO, 2020, there are two kinds of cases who will be classified as moderate. One is patients who are suspected to have COVID-pneumonia after presenting with cough or shortness of breath. These patients should have normal saturation levels on room air. The other ones are patients who present with mild infection but with exacerbation of co-morbidities such as diabetic ketosis (DKA), acute exacerbation of asthma or DKA.

#### Severe cases

Patients with COVID pneumonia or exacerbated comorbidity, who need oxygen support but not in any impending or already in respiratory failure. Patients can also be classified as severe if they have other severe medical complications such as but not limited to seizure, change in mentation or are in shock.

#### Critical cases

Patients who present with impending or are already in respiratory failure needing both oxygenation and mechanical ventilator support.

### Ethical Consideration

The study was approved by Institute of biotechnology, University of Gondar ethics and research committee; protocol number IOB/291/04/2021. Data was collected after permission was obtained. All the information obtained from the study participants were kept confidential.

### Statistical Analysis

SPSS statistical software package version 25.0 (SPSS Inc., Chicago, IL, USA) was used for statistical analysis. Chi-square test was used to determine association among categorical variables. The quantitative data were expressed as Mean ± SD and Median values. P value < 0.05 was considered as statistically significant.

## Results

### Socio-demographics characteristics and clinical features of study participants COVID-19

A total of 500 qRT-PCR confirmed COVID-19 patients were included in this study from a laboratory database. The median age of the study participants was 55.58±7.070 years, from these 338 (67.6%) of patients were males. 60 (12%) study participants were within critical disease and 298 (59.6%) with severe disease. Most of the study participants (350 (70%)) were older than 55 years. The median age was significantly higher in critical (ICU patients) group compared to moderate and severe groups. The duration of symptoms before admission was also higher in severe patients (Table 1).

**Table 1:** Socio-demographic characteristics and clinical features of study participants at MCCC, Addis Ababa, Ethiopia, 2021

| Variables | Category of variables | Frequency | Percent (%) |
| --- | --- | --- | --- |
| Mean Age | | $55.58 \pm 7.707$ | |
| Age | $<55$ | 150 | 30 |
| | $\geq 55$ | 350 | 70 |
| Gender | Male | 338 | 67.6 |
|  | Female | 162 | 32.4 |
| Disease Severity | Mild | 58 | 11.6 |
|  | Moderate | 84 | 16.8 |
|  | Severe | 298 | 59.6 |
|  | Critical | 60 | 12 |
| Comorbid | No comorbidity | 269 | 53.8 |
|  | Diabetics (DM) | 61 | 12.2 |
|  | Hypertension (HTN) | 58 | 11.6 |
|  | Asthma | 10 | 2 |
|  | Cardiac | 8 | 1.6 |
|  | HTN+DM | 55 | 11 |
|  | HTN + Asthma | 3 | 0.6 |
|  | HTN +DM +Cardiac | 14 | 2.8 |
|  | ARVI | 10 | 2 |
|  | COPD | 4 | 0.8 |
|  | CLD +DM+CHF | 8 | 1.6 |
| Patient outcome | Improved | 426 | 85.2 |
|  | Death | 74 | 14.8 |

### Liver, renal and cardiac function test results

Liver function test parameter values of overall study participants were elevated and three of these parameters such as AST, ALT and ALP were elevated among critical (ICU) patients (56.9±57.7, 58.5±63 and 114.6±60) respectively. All patients had an elevated Creatinine, whereas Urea was in the normal range renal function test parameters. Creatinine was elevated among severe patients than moderate cases, whereas urea was elevated among critical patients than moderate and severe patients. Troponin values of overall patients were elevated but CKMB were in normal range and troponin value markedly elevated among critical (ICU) patients. There was no marked difference between Moderate, severe and critical group for CKMB value (P value=0.03) (Table 2).

**Table 2:** Liver, cardiac and renal function test parameters among COVID-19 confirmed patients, at MCCTC, Addis Ababa, Ethiopia

| Parameters | Normal range | Overall<br>$\mu \pm SD$ | Moderate<br>$\mu \pm SD$ | Severe<br>$\mu \pm SD$ | Critical<br>$\mu \pm SD$ | P-value |
| --- | --- | --- | --- | --- | --- | --- |
| ALT,U/L | 0-33 U/L | $57 \pm 65.4$ | $67.2 \pm 89.4$ | $42.28 \pm 23.7$ | $56.9 \pm 57.7$ | 0.06 |
| AST,U/L | 10-35U/L | $53 \pm 55$ | $51.35 \pm 53$ | $41.4 \pm 28.48$ | $58.5 \pm 63$ | 0.028 |
| ALP,U/L | 45-87U/L | $104 \pm 58$ | $90.5 \pm 36.53$ | $103.4 \pm 73.7$ | $114.6 \pm 60$ | 0.092 |
| Creatinine mg/dl | 0.5-0.9 mg/dl | 1.37±6.3 | 0.95±0.75 | 3.13±14.36 | 0.96±0.74 | 0.043 |
| Urea mg/dl | 10-45mg/dl | 44.8±28.7 | 45±29.45 | 37.7±22.1 | 47±30 | 0.058 |
| Troponin pg/ml | 0-14pg/ml | 60.1±123.5 | 36.3±89.65 | 35.7±47.9 | 83±153 | 0.0356 |
| CK-MB U/L | 0-25U/L | 3.3±6.7 | 4.8±11.07 | 1.39±0.54 | 2.85±2.93 | 0.003 |

Elevated AST 76/154(49%), ALT 96/154(62.3%), creatinine (57/154), troponin (91/154) was seen among study subjects older than 55 years. 66.8% males, 65% ICU, had an elevated serum value of ALT and AST respectively. Troponin was found elevated among males (54%) and 59% among ICU (critical) patients (Table 3).

**Table 3:** Frequency of Liver and Renal function profiles in relation to normal range among COVID-19 patients admitted at MCCC, Addis Ababa, Ethiopia, 2021

| Parameters |  | Age |  | Sex |  | Disease severity |  |  |
| --- | --- | --- | --- | --- | --- | --- | --- | --- |
|  |  | <55n (%) | >55n (%) | Female n (%) | Male n (%) | Moderate n (%) | Severe n (%) | Critical n (%) |
| ALT,U/L | Elevated | 37(56) | 96(62.3) | 28(44.4) | 105(66.8) | 37(53.7) | 26(59) | 70(65.4) |
| AST,U/L | Elevated | 32(48.480) | 76(49.3) | 24(38) | 84(53.5) | 33(47.8) | 14(20.2) | 61(57) |
| ALP,U/L | Elevated | 20(30) | 42(27.2) | 17(27) | 45(28.7) | 14(20.2) | 15(21.7) | 33(30.8) |
| Creatinine mg/dl | Elevated | 13(19.6) | 57(37) | 11(17.4) | 59(37.57) | 22(31.8) | 9(13) | 39(36.4) |
| Urea mg/dl | Elevated | 11(16.6) | 62(40.2) | 15(23.8) | 58(36.9) | 23(33.3) | 9(13) | 41(38.3) |
| Troponin pg/ml | Elevated | 25(37.8) | 91(59) | 30(47.6) | 86(54.7) | 27(39.13) | 25(36.2) | 64(59.8) |
| CK-MB U/L | Elevated | 20(30.3) | 43(28) | 22(34.9) | 41(26.1) | 10(14.4) | 19(27.5) | 34(31.7) |

## Discussion

This study investigates the profiles of clinical chemistry tests of COVID-19 qRT-PCR confirmed hospitalized cases using routine clinical chemistry automated analyzer. The laboratory parameter values indicated that for COVID-19-positive cases, from routine clinical chemistry tests, such as liver function tests, like ALT, AST and ALP, Renal function tests, like Creatinine and Urea and Cardiac Function tests, Troponin(TnT) and ALP were found increased.

Abnormal liver function test results were seen in up to more than half of patients. 59% & 65.4% elevated ALT was found among critical and severe patients, 34.1% and 57% elevated AST among severe and critical patient was found respectively. Different studies showed that COVID-19 stratification based on disease severity, including the need for ICU admission and the extent of respiratory distress, indicated that serum AST and ALT were elevated in patients with critical COVID-19 compared to those with severe and moderate disease [1, 2, 27-30] and during the 2002-2004 SARS outbreaks this was also reported [31]. In COVID-19 the prognostic value of abnormal LFTs is not well defined but some studies states that abnormal LFTs, particularly elevated AST total bilirubin and ALT, are associated with increased disease severity,[1, 2, 20, 22, 27-30, 32] whereas other studies report as there is no association with disease severity and progression[33, 34]. Even though the mechanism of extra-pulmonary spread of SARS-CoV-2, whether hematogenous, Virus-mediated direct tissue damage or otherwise, remains elusive but different reports across the globe suggest that SARS CoV-2, migration in the blood, intestines may infect the liver and damage hepatocytes, which could result AST, Bilirubin and ALT values elevation[35-37].

Secondary liver damage due to the administration of hepatotoxic drugs, respiratory distress syndrome-induced hypoxia, systemic inflammatory response, and multi organ failure are believed to be associated risk factors for COVID-19-related liver dysfunction [38, 39].

Among renal function tests, serum urea and Creatinine showed elevated value among severe (56.8%, 20.45%) and critical (59.8%, 36.4%). Old age study participants had elevated serum urea (40.2%), Creatinine (37%). Similar studies revealed that renal function marker alterations in COVID-19 evidenced by increased serum urea and Creatinine values. Kidney tubular cells, which express the ACE2 receptor on their cellular surface, could be directly infected by SARS COV-2[19, 21]. Evidences also revealed that kidney-resident cells could interact with circulating mediators resulting in microcirculatory derangement, endothelial dysfunction, and tubular injury [8, 15, 40].

Reports indicate that about 25–30% of individuals infected with SARS COV-2 develop acute kidney injury (AKI) and this was associated with increased mortality risk [15, 41, 42]. AKI occurred at much higher rates in critically ill patients admitted to hospitals, ranging from 78% to 90% and it is a frequent complication of COVID-19 which is associated with mortality [43-45]. Cardiac injury is a common clinical feature of COVID-19 patients; this could result from SARS-CoV-2 infection as a result of direct and indirect effect on cardio myocytes, including acute myocardial infarction, impaired renal function, heart failure, arrhythmias, myocarditis, sepsis, septic shock, cardiac arrest and pulmonary embolism [23, 25, 26]. In cardiac markers, CK-MB and troponin serum value was found elevated among critical (59.8%), Severe (56%) and Moderate (39%) patients. A greater frequency and magnitude of troponin elevations in hospitalized patients has been associated with more-severe disease.

Petrosillo et al., which compares the AST enzymes of different corona viruses, found that an average elevated amount up to 31.5% in SARS COV-2 cases than the others [46]. This elevation of AST enzymes could be associated either with cardiac problems associated with COVID-19, where AST together with CK-MB, LDH and troponin could be high. Underlying cardiac disease was reported as among the major risk factors in susceptible to COVID-19 [24, 47, 48]. Myocardial injury, with elevation of cardiac biomarkers above the 99th percentile of the upper reference limit, occurred in 20–30% of hospitalized patients with COVID-19, with higher rates (55%) among those with pre-existing cardiovascular disease [49-51].

Guo *et al*. studied 187 COVID-19 patients, of whom 52 (27.8%) had a myocardial injury as determined by elevated levels of troponin [52]. The incidence of cardiac injury ranged anywhere between 8 and 12% in various studies; the incidence being 13-fold higher in the ICU/severe category [30, 53, 54]. Moreover, the patients admitted to the ICU had a 2.2-fold higher troponin level when compared with the non-ICU [53, 54]. Therefore, cardiac damage biomarkers evaluation upon admission and the longitudinal monitoring during hospital stay helps to prompt intervention in order to improve the progression of the disease and could represent a significant tool for the early detection of cardiac injury [48, 55].

## Conclusions

Abnormal serum values of organ function biomarkers have been associated with COVID-19 disease severity and a worse prognosis. Organ function biomarkers help for risk stratification and for predicting of COVID-19 disease severity in order to guide clinical care. Among all, AST, ALT, Creatinine, urea, Troponin and CKMB represent the most predictive parameters of organ failures among critical COVID-19. Organ function markers should be monitored regularly for the management of COVID-19.

## Data Availability

all data can be available up on request

## Ethical approval

The study protocol was reviewed and approved by University of Gondar, Institute of Biotechnology Ethical Committee with protocol approval number IoB/291/04/2021. Permission was obtained from St. Paul Medical Millennium Medical College. Written informed consent were obtained from all participants and signed prior to the commencement of the interview and it was in accordance with the principles of the Helsinki II declaration. All the information obtained from the study participants were kept confidential

## Acknowledgment

The authors would like to thank the University of Gondar, Institute of Biotechnology for funding and facilitating the PhD research work of Yakob Geberegzabher Tsegaye. Defense University, college of health science also thanked for providing scholarship for the PhD student. We are also very grateful for the Millennium COVID-19 Care and Treatment Center (MCCTC) for providing facilities to meet the COVID-19 Patients.

## Availability of data and material

All the available data were included in the manuscript.

## Funding

This manuscript is emanated from the PhD research project was supported by the UoG, Institute of Biotechnology

## Author’s contribution

YGT, MB, TW and NB conceptualize this research work, YGT,, MB, TW and NB designed the methodology and data analysis part of this manuscript, YGT, MB, TW, AA, MA, MG and NB Write, Review and edit this manuscript.

## Conflict of interest

The authors declare that they have no conflict of interest.

## Notes

### Competing Interest Statement

The authors have declared no competing interest.

### Funding Statement

This is the PhD thesis work of Yakob Tsegaye and it was funded by university of Gondor.

### Author Declarations

The study was approved by Institute of biotechnology, University of Gondar ethics and research committee; protocol number IOB/291/04/2021.

